# Functional connectivity of the nucleus accumbens predicts clinical course in treated and non-responder adult ADHD

**DOI:** 10.1101/2024.01.04.24300820

**Authors:** Ahmed Zaher, Jan Leonards, Andreas Reif, Oliver Grimm

## Abstract

Attention deficit hyperactivity disorder (ADHD) is a neurodevelopmental condition that often persists into adulthood, contributing to a challenging trajectory characterised by increased rates of psychiatric comorbidity. Contrary to previous assumptions, the clinical course of ADHD in adulthood is characterised by dynamic changes influenced by stimulant treatment. To improve the precision of treatment, the identification of a predictive neuroimaging biomarker would be of great clinical importance.

In this study, we explore the potential of seed-based functional connectivity (FC) as a predictive tool for assessing the clinical course of ADHD symptoms. We conducted a longitudinal follow-up study of 54 adult ADHD patients who underwent magnetic resonance imaging (MRI) using a 3 Tesla scanner. All patients received stimulant treatment during an initial run-in period. After an average of three years, only subjective responders continued treatment (n=34), while subjective non-responders discontinued medication (n=20). We reassessed patients after three years of treatment to 1) evaluate the prediction of individual outcome by baseline fMRI and 2) to investigate differences in prediction by baseline fMRI depending on long-term treatment vs. discontinuation.

Our investigation focused on determining whether a relationship could be established between resting-state FC of the nucleus accumbens (NAc) and the trajectory of symptom development, as well as identifying possible differences in FC between individuals who had been on long-term stimulant treatment and those who had not used stimulants at the time of follow-up.

In general, reduced FC between the NAc and the default mode network (DMN) was associated with initially higher symptom burden, whereas patterns of improvement correlated with reduced FC between the NAc and the salience network (SN).

Comparatively, higher FC between the NAc and the SN was associated with better symptom outcomes in patients receiving long-term stimulant medication, whereas lower FC between the NAc and the SN was correlated with a more favourable prognosis in non-responders.

Our prospective fMRI study suggests that functional connectivity between the dopaminergic reward circuitry and the SN, especially the insula, is predictive of better ADHD symptom outcomes. This work highlights the potential of dopaminergic functional connectivity as a prognostic factor in ADHD. While future research with larger samples and longer follow-up is warranted, our study highlights the role of the NAc in the prognosis of ADHD.

## 1. Introduction

Attention deficit hyperactivity disorder (ADHD) stands as a prevalent neurodevelopmental condition characterized by inattentiveness, impulsivity, and hyperactivity. These traits typically emerge in childhood and often persist into adulthood. Globally, the prevalence of ADHD in children is estimated to be between 5% to 7% (Spencer et al., 2007), while among adults, it is reported to be 3% (Franke et al., 2018). Importantly, ADHD is associated with an elevated risk of comorbid conditions across the lifespan of individuals affected by it. In both childhood and adulthood, ADHD patients experience a diverse range of psychiatric comorbidities, including but not limited to, substance use disorders (SUD), depression, as well as diminished quality of life, reduced treatment effectiveness, and even increased mortality when compared to the general population (Franke et al., 2018; Matthies et al., 2018; Weibel et al., 2020).

Despite being established as a neurodevelopmental disorder, the underlying mechanisms of ADHD neuropathology in the involved brain networks remain partially blurred. One pivotal brain network implicated in both ADHD and other psychiatric disorders is the reward network (Grimm et al., 2021). Numerous studies have consistently highlighted altered neural activity and functional connectivity (FC) patterns in reward circuits among individuals suffering from ADHD when contrasted with neurotypical subjects (Grimm et al., 2021; Plichta and Scheres, 2014; von Rhein et al., 2015). In particular, the nucleus accumbens (NAc) has been shown to be a central player in the neurobiology of ADHD symptoms (Hoogman et al., 2017; Shaw et al., 2014; Sonuga-Barke, 2005).

The multifaceted conglomerate of symptomatology representing ADHD changes over the course of time exhibiting different functional connectivity patterns that even shows a sex specific difference (Dupont et al., 2022).

Our study investigated whether resting-state FC patterns of the nucleus accumbens (NAc) could predict the course of ADHD symptoms in newly diagnosed adult patients. After conducting baseline f-MRT scans and comprehensive clinical evaluation at the point of diagnosis (*t_1_*), we conducted a clinical follow-up after approximately three years later (*t_2_*). We measured the burden of ADHD symptoms using the Wender-Reimherr-Interview (WRI) which was also used at (*t_1_*) to diagnose patients. Change in WRI scores over time was calculated (Δ WRI: WRI *t_2_* - WRI *t_1_*) and then correlated with resting-state fMRI brain scans obtained at *t_1_* to establish a connection between clinical scale scores and NAc-FC patterns. We additionally explored the influence of medication on FC patterns, as we observed that around 30% of the participants had discontinued stimulants medication at the time of follow-up.

**Figure 1:**
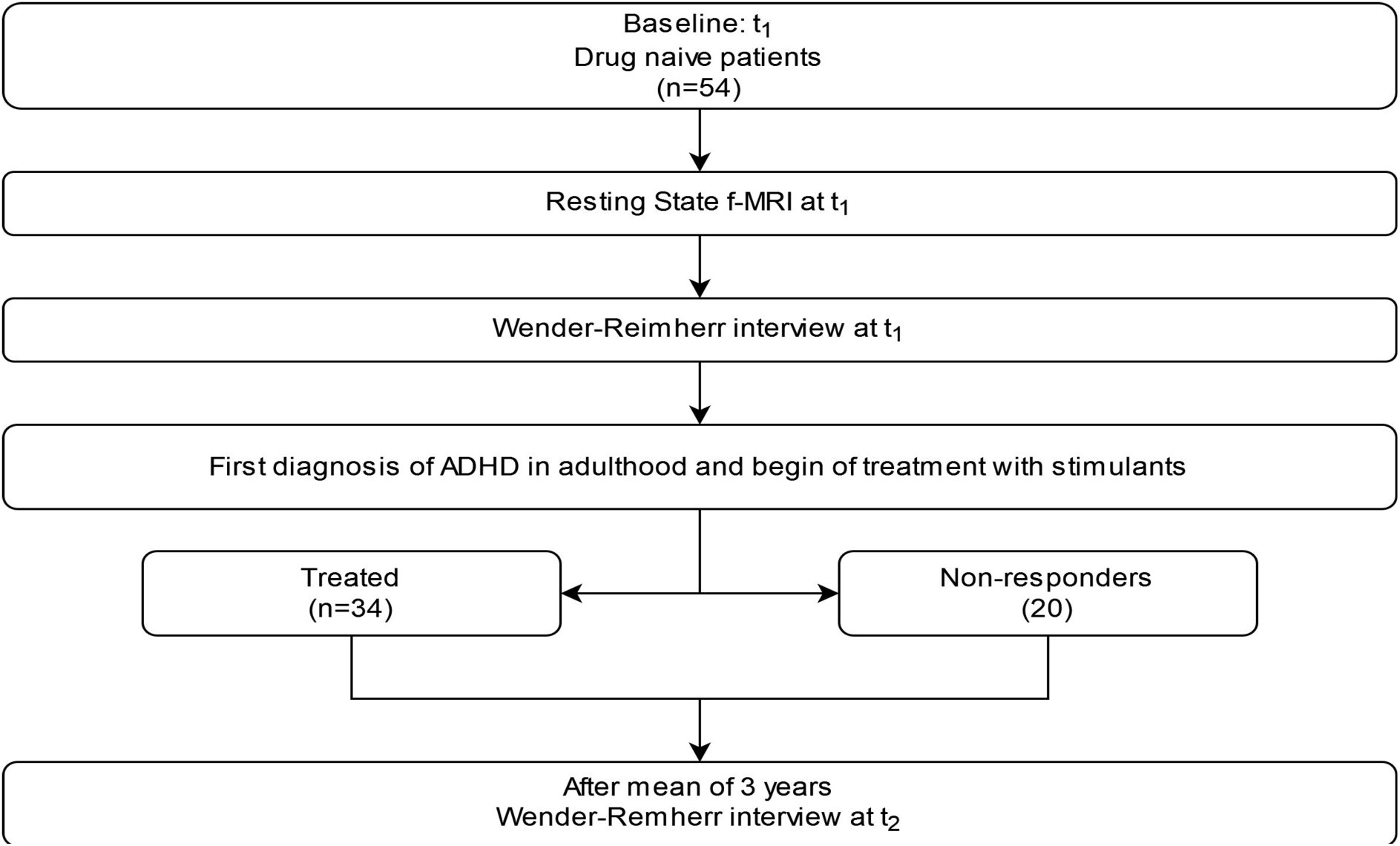
Study Workflow.

## 2. Methods

### 2.1 Participants

The study includes n=54 physically healthy ADHD patients who received their diagnosis in adulthood at *t_1_* (for demographics see table 1). Recruitment took place at the University Hospital; Goethe University Frankfurt am Main. Inclusion criteria were 1) age between 18 and 50 years; 2) sufficient understanding of the German language and, 3) established ADHD diagnosis on the basis of the DSM-IV criteria. Exclusion criteria were other mental illnesses (apart from ADHD, depression and SUD), serious acute or chronic physical diseases, pregnancy, as well as exclusion criteria of the MRI examination.

**Table 1:** Demographics (participants, n=54); M= mean.

| | treated $t_2$ ($n=34$) | non-responder $t_2$ ($n=20$) | statistics | p |
| --- | --- | --- | --- | --- |
| Age, M in years | 30.9 | 30.1 | $F=0.113$ | 0.738 |
| Female | 14 (25.9) | 9 (16.7) | $\chi^2 < 0.01$ | 0.975 |
| Male, n (%) | 19 (35.2) | 12 (22.2) |  |  |
| $\Delta$ WRI ($t_2-t_1$), M | -10.6 | -5.10 | $F=12.2$ | 0.001 |

The approval to conduct the study was given by the local ethics commission (Faculty of Medicine, University Hospital, Goethe University, Frankfurt am Main) and is subject to the Declaration of Helsinki of the “World Medical Association: Ethical Principles for Medical Research Involving Human Subjects” and the “Guidelines for Good Clinical Practices (GCP)”. In addition, the study was registered as a clinical trial in the German study registry under the ID: DRKS00011209. Written informed consent was obtained from each volunteer before the start of the study.

At *t_1_* the diagnosis was established using the Wender Reimherr Interview (WRI) that measures the severity of ADHD symptoms using seven subscales and an overall score indicating the severity of the symptoms. After fulfilling the revised DSM-IV diagnosis criteria for ADHD in adulthood all subjects were prescribed methylphenidate. After a mean period of 6 months a group of 20 subjects did not respond to medication and stopped the medication.

After a mean period of 3 years: *t_2_*, all subjects were contacted and we repeated the WRI. Participants were asked at *t_2_* if they were still taking treatment for their ADHD-Symptoms or not. 34 patients were still taking stimulants (Methylphenidate; treated, n= 34) while the remaining 20 were no longer taking medication at *t_2_*; (non-responder, n=20) at *t_2_*.

### 2.2 The Wender-Reimherr-Interview (*WRI)*

A trained and results-blinded registered psychiatrist conducted the German version of the Wender-Reimherr-Interview (WRI) (Rösler et al., 2021) to assess ADHD-specific symptoms in study participants.

The WRI is a structured psychopathological interview that assesses the severity of ADHD symptoms in seven categories including: attention difficulties, hyperactivity, temperament, affective lability, emotional reaction to stress, disorganization, and impulsivity. Each category is assigned a score depending on the severity of the symptom. An overall global score is calculated based on the seven categories. We used the individual patients’ global scale score in our statistical analysis.

We calculated the change of scores over time (Δ WRI: *t_2_-t_1_*) by subtracting the global WRI-score of the first assessment (WRI *t_1_*) from that of the second assessment (WRI *t_2_*).

## 3. Data analysis: fMRI

### 3.1 fMRI acquisition

For the MRI measurements in our study, we used a 3 Tesla full-body MR scanner (Siemens Magnetom Trio syngo MR A35) at the Brain Imaging Center in Frankfurt am Main. An eight-channel head coil was also utilized for data acquisition. We acquired a t1-weighted sequence (MPRAGE) with a duration of 4:28 minutes, as well as a gradient echo sequence for functional imaging data, which lasted 8:01 minutes. The sequence parameters for the MPRAGE sequence were as follows: repetition time (TR) = 1900 ms, echo time (TE) = 3.04 ms, TI = 900 ms, flip angle = 9, field of view (FoV) = 256 x 256 mm, and voxel size = 1 x 1 x 1 mm. For the EPI sequence, the parameters were as follows: TR = 1800 ms, TE = 30 ms, flip angle = 90, FoV = 192 x 192 mm, 28 layers with 4 mm thickness, and voxel size = 3 x 3 x 4 mm. To minimise head movement during the MRI scans, we used foam pads for the study participants.

### 3.2 Pre-processing

To reduce variability in the blood-oxygen-level dependent (BOLD) signal of the images, we used the preprocessing algorithm of the SPM12-based CONN-toolbox V18.b. The images of the participants were first realigned and slice-time corrected. They were then normalised to the Montreal Neurological Institute (MNI) template, resampled to 3mm isotropic voxels, and smoothed with an 8mm full-width at half maximum Gaussian kernel. The CONN toolbox’ default denoising pipeline was applied to further minimise noise in the data, including band-pass filtering in the 0.01-0.1Hz frequency range to remove non-neural signals, regression of motion parameters and their first-order derivatives, and correction for signals from cerebrospinal fluid and white matter using the a CompCor strategy (Behzadi et al., 2007)

### 3.3 Definition of region-of-interest masks

In our study, we used a seed-based connectivity analysis based on subcortical region-of-interest (ROI) seeds. The NAc was selected from brain regions that are central to the mesolimbic brain reward systems, as previously defined in the OTI Atlas (Pauli et al., 2018). This atlas was constructed using high-spatial resolution t1-and t2-weighted structural images, with tissue boundaries used to delineate subcortical nuclei, which were then combined to form a probabilistic atlas. From this atlas, we selected the NAc as our seed ROI for the connectivity analysis.

### 3.4 Data analysis: group statistics

Data analysis was done with the CONN toolbox v18.b and SPM12. The workflow for striatal seeds has been described before (Hamzehpour et al., 2023) We extracted the residual BOLD-time course from the seed ROI (NAc) and correlated it with other voxels in the brain in order to calculate the first-level correlation maps. We then transformed the first-level correlation coefficient maps into a normally distributed z-score. The transformed maps were then used as the basis for multiple regression tests and 2×2 between-subjects ANOVA interaction. In a first step we performed a regression test to calculate a possible relationship between the initial symptom load WRI *t_1_* and the FC of NAc and in a second analysis the change in symptom load over time (Δ WRI) in all patients (n=54). Age and sex were included as covariates in the model to account for age and sex-related variability between the subgroups of interest.

To explore whether the neural basis of connectivity patterns differed between treated and non-responder patients at *t_2_*, we used a 2×2 ANOVA test in our third step to study the interaction of medication separately. For correction of multiple testing during second-level statistics we used cluster-wise whole-brain analysis which uses a combination of an uncorrected p<0.001 height threshold to initially define clusters of interest from the original statistical parametric maps, and an FDR - corrected p<0.05 cluster-level threshold to select the significant clusters among the resulting clusters.

## 4. Results

### 4.1 ADHD symptom severity

In order to measure the ADHD symptoms load, the global score of the WRI was calculated at the time of diagnosis and at the time of follow up. WRI *t_1_* was (M=14.5, SD=4.16, Min/Max= 6 / 22), WRI *t_2_* was (M=6.06, SD=5.55, Min/Max= 0 / 21). The Δ WRI showed (M=-8.46, SD=5.86, Min/Max= -21 / 0).

**Table 2.** Pearson’s Correlation between change of WRI scores (Δ WRI) over time as well as initial WRI scores (WRI t1) and receiving stimulant medication at *t2*. NB: *: p<0,01

|  | *Stimulants at $t_2$* | *$\Delta$ WRI* | *WRI $t_1$* |
| --- | --- | --- | --- |
| Stimulants at $t_2$ | 1 | -0.46* | -0.19 |
| $\Delta$ WRI | -0.46* | 1 | -0.42* |
| WRI $t_1$ | -0.19 | -0.42* | 1 |

### 4.2 ADHD symptoms and Functional Connectivity of the NAc

#### 4.2.1 ADHD symptoms at baseline and FC in the total sample

A significant correlation was observed between the severity of symptoms at the time of diagnosis (WRI *t_1_*) and the FC between the NAc and the right paracingulate gyrus, where a lower connectivity between the NAc and the paracingulate gyrus correlated with higher severity of ADHD symptoms in adulthood. (Cluster k=93; MIN Coordinates (x,y,z): 12, 42, 32; pFDR: 0.01; t:5.64)

**Figure 2.**
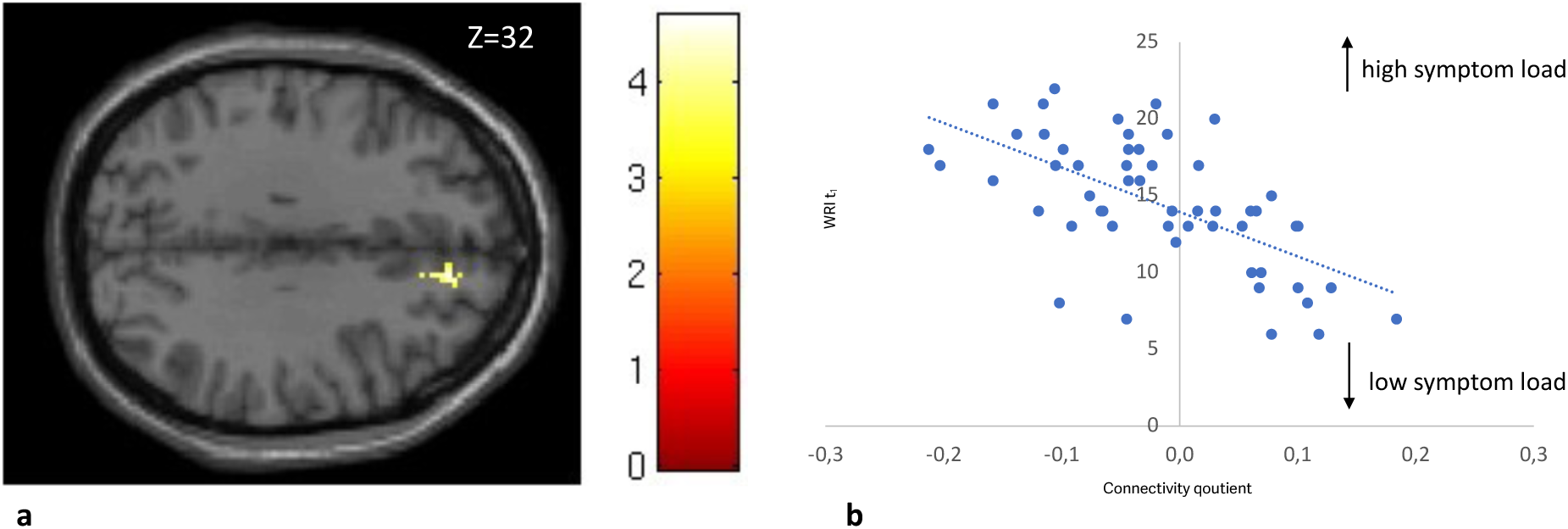
Correlation between global score WRI t1 and the FC-quotient between NAc and the paracingulate Gyrus right in all subjects. In 2A the most significant cluster of the connectivity analysis: the paracingulate gyrus right and its localization is demonstrated z=32, (Cluster k= 93; MIN-Coordinates (x,y,z) : 12, 42, 32 ; pFDR: 0.01 ; t:5.64). In 2B a scatterplot of individual FC-Quotient against global score WRI t1 of each subject is shown. FC: Functional connectivity; NAc: Nucleus accumbens; WRI: Wender Reimherr Interview

#### 4.2.2 Change in ADHD symptoms and FC in the total sample

A significant correlation was observed between the long-term change of the global score: Δ WRI (*t_2_-t_1_*) and the FC between the NAc and the insular cortex, where a lower connectivity between the NAC and the right insular cortex predicted a better outcome of the ADHD symptoms. (Cluster k= 101; MIN-Coordinates (x,y,z) : 36, 12, 4; pFDR: 0.01; t:5.33)

**Figure 3.**
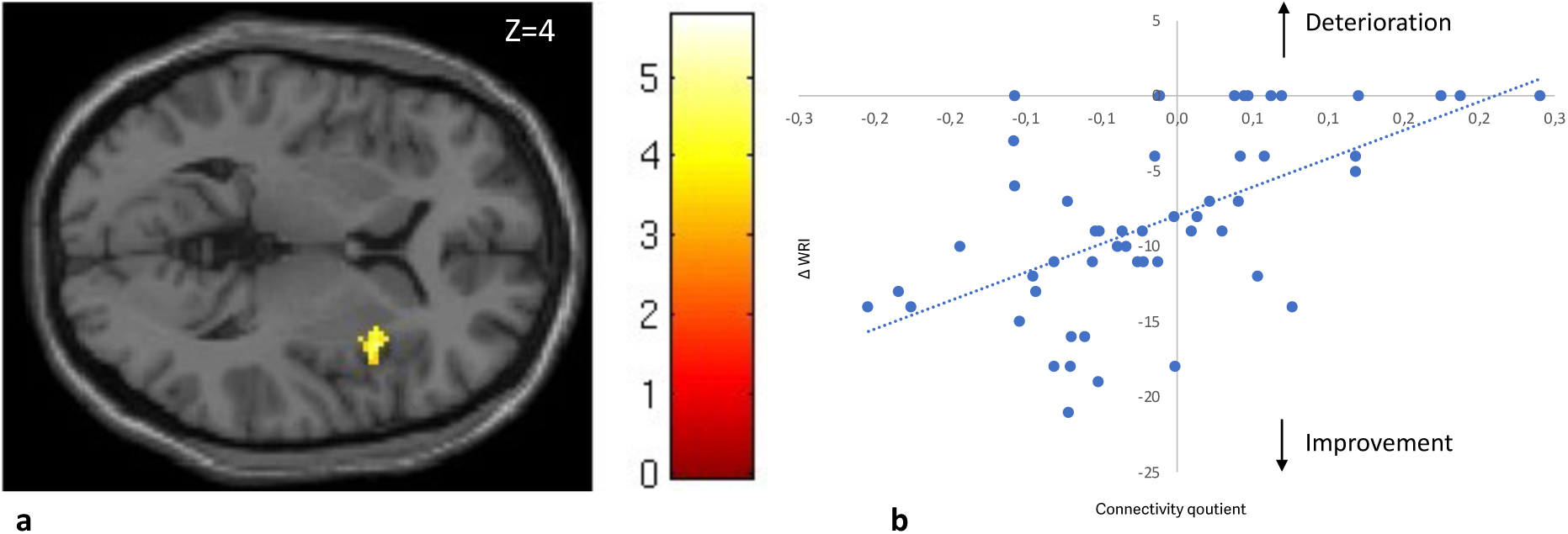
Correlation between Δ WRI (t2-t1) and the FC-quotient between the NAc and the anterior insula right in all subjects. In 3A the most significant cluster of the connectivity analysis: the anterior insula right and its localization is demonstrated z=4, (Cluster k=101; MIN-Coordinates (x,y,z) : 36, 12, 4 ; pFDR: 0.01 ; t:5.33) In 3B a scatterplot of individual FC-quotient against Δ WRI each subject is shown. NB: Arrow direction explains the development of symptoms over time. FC: Functional connectivity, NAc: Nucleus accumbens, WRI: Wender Reimherr Interview

#### 4.2.3 ADHD symptom severity in continued vs. discontinued treatment

Next, we compared the correlation of FC to Δ WRI in those receiving stimulants and those not at *t_2_*. In those receiving treatment, a higher FC between the NAC and the insular cortex predicted a better overall long-term outcome. Meanwhile we observed that in those not receiving medication at *t_2_* higher FC predicted a worse outcome, (Cluster k=91; MIN-Coordinates (x,y,z): 42, -12, -2; pFDR :0.04; t: 4.88)

**Figure 4.**
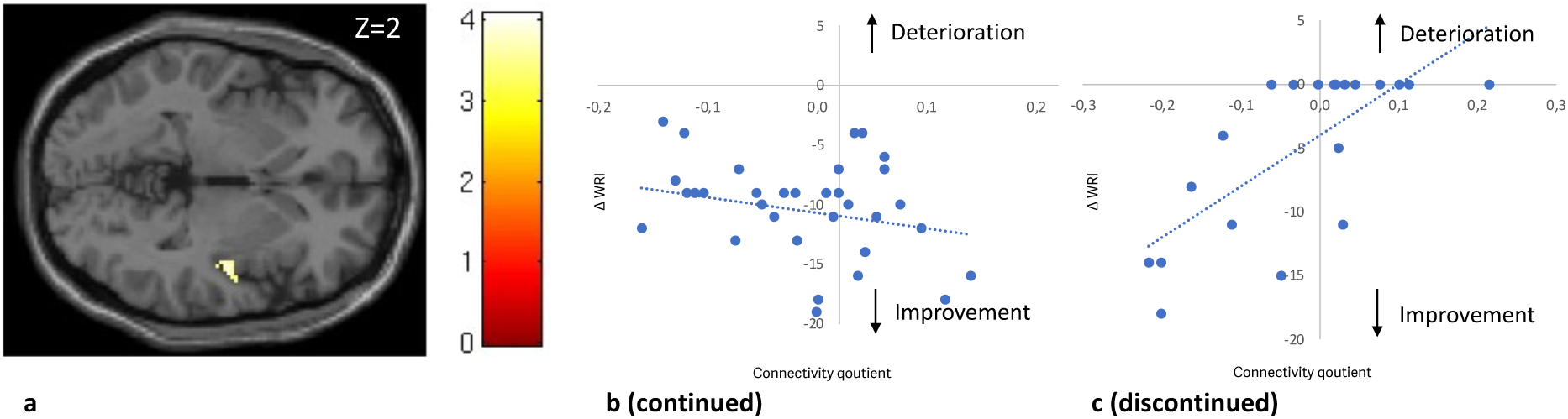
Correlation between Δ WRI (t2-t1) and the FC-quotient between NAc and the posterior insula in patients with continued vs discontinued treatment. In 4A the most significant cluster of the connectivity analysis: the posterior insula right and its localization is demonstrated z=-2, (Cluster k=91; MIN-Coordinates (x,y,z) : 42, -12, -2; pFDR :0.04; t: 4.88). In 4B a scatterplot of individual FC-quotient of patients with continued treatment against Δ WIR of those subjects is shown. In 4C a scatterplot of individual FC-quotient of patients with discontinued treatment against Δ WIR of those subjects is shown NB: Arrow direction explains the development of symptoms over time. FC: Functional connectivity, NAc: Nucleus accumbens, WRI: Wender Reimherr Interview

### 4.3 Movement-related effects

We extracted mean-motion and max-motion values from the treated vs. non-responder (n = 54) and performed independent samples tests. Treated and non-treated patients did not show significant motion changes (p > .05). We did not detect any differences between the two groups in mean-motion (p = 0.25) or max-motion (p = 0.55).

**Table 3:**
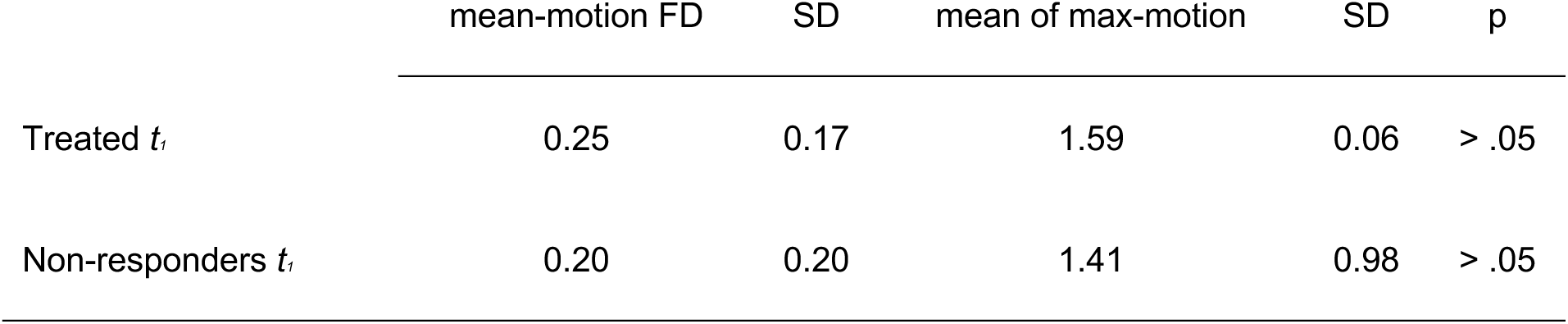
Movement related effects in all treated and non-responder subjects (participants, n=54; treated, n=34; non-responder, n=20)

## 5. Discussion

Our study found that changes in brain connectivity between the NAc and the paracingulate cortex (PCC) and insular cortex (IC) predicted symptom improvement between the two time points *t_1_* and *t_2_*. The direction of FC patterns differs depending on whether medication is continued or discontinued due to treatment failure.

First, analysis showed that a higher symptom load at *t_1_* correlated with decreased FC between the NAc and the right paracingulate gyrus: an important region of the default mode network of the brain (DMN), reflecting previous reports that ADHD patients exhibit hypo-connectivity between the reward system and the DMN (Castellanos et al., 2008; Sutcubasi et al., 2020; Tomasi and Volkow, 2012). Improvement in symptoms over time was associated with decreased FC between the NAc and the right anterior insular cortex (aIC) of the salience network (SN).

Second, when considering the medication status of subjects at *t_2_*, a significant correlation emerged between receiving stimulant medication and improvement in global symptom scores. Dividing our subjects into two groups based on their medication status at *t_2_*, we found in the treated group, a higher FC between the NAc and the right posterior insular cortex (pIC) correlated significantly with improvement under medication. Conversely, improvement in the non-responder group correlated with lower FC between the NAc and the right pIC.

These findings have to be interpreted from the background of previously described alterations in the reward system in ADHD (Cao et al., 2009; Castellanos et al., 2008; Volkow et al., 2007): The reward circuitry in humans, including the ventral tegmental area (VTA), the ventral striatum, and dopaminergic projections to the NAc, forms a crucial part of the brain’s reward system(Koob and Le Moal, 2008).In the pathophysiology of ADHD symptomatology, the brain reward circuit is considered to be one of the major brain hubs involved where changes in the functional connectivity of the NAc and the DMN have been implicated in the development of ADHD (Alves et al., 2019). In particular, deficits in the intra circuitry of the brain’s reward system and alterations in the connectivity of the NAc are observed in ADHD patients (Stark et al., 2011). Children suffering from ADHD have been recently shown to have altered brain network developmental trajectories between cortico-limbic regions and between visual and higher-order cognitive networks (Soman et al., 2023).

Our subjects displayed a significant correlation between higher ADHD symptom load and the FC of the NAc to the right paracingulate gyrus. Hypo-connectivity between these regions correlated positively with symptom load. The paracingulate gyrus, part of the cingulate cortex in the DMN, is active during rest and self-referential mental activities (Amunts et al., 2020). Prior studies have established structural connections between the NAc and the DMN, indicating a potential influence on ADHD symptomatology (Alves et al., 2019).The DMN is active during rest, involved in self-referential thoughts, and mind-wandering. Altered DMN activity could interfere with an individual’s ability to sustain attention. Interactions between the DMN and other brain networks, such as the SN, involve distinct topological structures that support reward processing and emotion regulation (Yankouskaya et al., 2022).

Our study shows that patterns of FC between these key areas can play a role in predicting the course of ADHD-symptoms. Generally decreased FC between the DMN and the NAc correlated with high symptom load.

In ADHD individuals, reduced FC between the DMN and the reward system suggests disruptions in cognitive control processes related to reward processing. The dorsal anterior insula (dAI), responsible for cognitive control, appears to be implicated in these disruptions.

The decreased FC between the reward system and the DMNsuggests that individuals with higher ADHD symptom load experience disruptions in their ability to effectively control cognitive processes related to reward processing.

Apart from altered connections to the DMN and dACC, ADHD patients’ symptom improvement over time, as measured by the change in WRI, was significantly linked to reduced functional connectivity between the NAc and the anterior insular cortex (aIC).

Located in the lateral sulcus separating the frontal and temporal lobes, the insular cortex is a complex, highly connected cortical hub. It plays a critical role in interoception, self-awareness, and emotional regulation (Benarroch, 2019), and is key in managing attention through its interactions with the fronto-parietal network (FPN) and the DMN (Menon, 2011; Menon and Uddin, 2010). As a central structure in the SN, it’s pivotal in transitioning between the DMN and other networks, assessing the relevance of sensory information and mental events (Seeley et al., 2007; Sheline et al., 2009). Notably, adults with persistent ADHD symptoms exhibit reduced insular density, especially thinning of the left insular grey matter in combined subtype ADHD (Maier et al., 2023). The observed FC patterns and their relationship to ADHD symptom changes can be partly explained by the ventral anterior insula (vAI) subdivision, linked to affective processes. Increased FC between the reward system and the anterior insula in individuals with unimproved or worsening symptoms may suggest amplified affective responses to rewards, potentially exacerbating symptoms. Conversely, symptom improvement associated with reduced FC might indicate normalized affective responses. Upon examining medication status, we found a significant correlation between receiving stimulant treatment at the second time point (t_2_) and symptom improvement, as indicated by the WRI at *t_2_*. This suggests a relationship between symptom amelioration and stimulant medication. Patients who responded positively to stimulant medication at *t_2_* exhibited a pattern where increased functional connectivity (FC) between the NAc and the posterior insular cortex (PI) corresponded with WRI-scale improvement. In contrast, non-responders, who did not improve and were not on medication at *t_2_*, showed an opposite pattern, where increased FC was linked with symptom deterioration. This pattern suggests that in ADHD individuals, the PI might emit stronger, possibly dysregulated interoceptive signals, contributing to symptom complexity.

Medication, by influencing the connectivity between the PI and the reward system, may help regulate these interoceptive signals (or urges) and mitigate impulsive responses. This interpretation aligns with the concept that the insula’s original role in urge processing was evolutionarily geared toward satisfying primary biological needs. Stimulants like methylphenidate (MPH) or amphetamines can modulate connectivity patterns in the brain, affecting neural networks associated with attention and impulse control. The differences in connectivity changes between the treated and non-responder groups may be due to the mechanisms of stimulant treatment. Wong et al. showed that stimulant drugs had an effect on the FC of fronto-parietal (FPN) brain networks. (Wong and Stevens, 2012). Picon et al found that ADHD patients exhibited increased connectivity within the DMN, specifically between the Posterior Cingulate Cortex (PCC) and Lateral Parietal Cortex (LLP) after methylphenidate treatment (Picon et al., 2020). However, this likely reflects the prediction independent of the medication continuation status.

Although at a group level MPH has been proven to be effective in ameliorating symptoms, there is individual variability in response, which affects clinical outcomes as shown by the two groups. Further understanding of the effects of stimulants on an individual level is crucial to interpreting the differences between the treated and the non-responder group. It’s possible that in non-responders, higher connectivity reflects a maladaptive response to medication withdrawal, leading to symptom worsening. Some limitations need to be addressed: While we describe NAc connectivity patterns, we are aware that rs-fMRI in our technical setting might not prove optimal to reliably delineate different brain nuclei. In addition, future studies might use more sophisticated surface-based analysis and a more fine-grained analysis of the anterior-posterior shift in the insula’s functional subdivision (Chang et al., 2013). Furthermore, resting state fMRI is not directly related to behavioral outcomes. Future research should supplement our findings with task-based behavioral paradigms related to ADHD pathophysiology, such as reward anticipation. Furthermore, our sample size is small. The results need to be replicated in a larger sample, so in future studies; we will aim for a larger sample size and thus report the smaller sample size as a limitation. Future studies should also include children and adolescents to observe the development trajectory of the reward system FC to the DMN and SN.

In summary, our study found that FC patterns between the reward system and the DMN and SN correlate with the clinical course of ADHD symptoms and the response to medication. Further studying of the reward system and its role in mediating between both networks is of importance especially in ADHD patients. FC of the reward system to DMN and SN has the potential to be used as a biological marker of response to stimulants-medication and clinical prognosis of ADHD. In conclusion, the correlation between higher NAc-connectivity and divergent treatment outcomes in ADHD patients after three years is a complex phenomenon. The interplay of clinical severity, treatment mechanisms, neuroplasticity, individual variability, and the length of follow-up may all contribute to these findings. Further research, possibly with extended follow-up periods, randomization, and more comprehensive assessments, is needed to better understand the underlying reward system mechanisms and implications for clinical practice.

## Data Availability

All data produced in the present study are available upon reasonable request to the authors

## Author disclosure

Financial support for this study was received from the European Union’s Horizon 2020 research and innovation program under grant agreement 667302: Comorbid Conditions of ADHD (CoCA) as well as the Deutsche Forschungsgemeinschaft (DFG grant 445498183, Oliver Grimm). The authors are solely responsible for the content of this publication. The EU had no further role in study design; in the collection, analysis and interpretation of data; in the writing of the report; and in the decision to submit the paper for publication.

OG and AR conceptualized the research; AZ, JL and OG collected the data. AR performed data analysis. AZ, JL, AR and OG designed the manuscript. AZ and OG wrote the manuscript. All authors revised the manuscript and approved the final version of the manuscript for publication.

AR received speaker and consultation fees from Boehringer Ingelheim GmbH COMPASS, Janssen Pharmaceuticals, LivaNova USA, Inc., Medice, Sage Therapeutics, Inc., and Shire within the last three years.

OG received speaker and consultation fees from Boehringer-Ingelheim and Takeda within the last three years.

AZ and JL report no conflicts of interest.

## Bibliography

Alves, P.N., Foulon, C., Karolis, V., Bzdok, D., Margulies, D.S., Volle, E., Thiebaut de Schotten, M., 2019. An improved neuroanatomical model of the default-mode network reconciles previous neuroimaging and neuropathological findings. Commun Biol 2, 370. 10.1038/s42003-019-0611-3

Amunts, K., Mohlberg, H., Bludau, S., Zilles, K., 2020. Julich-Brain: A 3D probabilistic atlas of the human brain’s cytoarchitecture. Science (1979) 369, 988–992. 10.1126/science.abb4588

Behzadi, Y., Restom, K., Liau, J., Liu, T.T., 2007. A component based noise correction method (CompCor) for BOLD and perfusion based fMRI. Neuroimage 37, 90–101. 10.1016/j.neuroimage.2007.04.042

Benarroch, E.E., 2019. Insular cortex. Neurology 93, 932–938. 10.1212/WNL.0000000000008525

Cao, X., Cao, Q., Long, X., Sun, L., Sui, M., Zhu, C., Zuo, X., Zang, Y., Wang, Y., 2009. Abnormal resting-state functional connectivity patterns of the putamen in medication-naïve children with attention deficit hyperactivity disorder. Brain Res 1303, 195–206. 10.1016/j.brainres.2009.08.029

Castellanos, F.X., Margulies, D.S., Kelly, C., Uddin, L.Q., Ghaffari, M., Kirsch, A., Shaw, D., Shehzad, Z., Di Martino, A., Biswal, B., Sonuga-Barke, E.J.S., Rotrosen, J., Adler, L.A., Milham, M.P., 2008. Cingulate-Precuneus Interactions: A New Locus of Dysfunction in Adult Attention-Deficit/Hyperactivity Disorder. Biol Psychiatry 63, 332–337. 10.1016/j.biopsych.2007.06.025

Chang, L. J., Yarkoni, T., Khaw, M. W., Sanfey, A. G., 2013. Decoding the Role of the Insula in Human Cognition: Functional Parcellation and Large-Scale Reverse Inference. Cerebral Cortex 23, 739–749. 10.1093/cercor/bhs065

Chiang, H.-L., Tseng, W.-Y. I., Wey, H., Gau, S., 2022. Shared intrinsic functional connectivity alterations as a familial risk marker for ADHD: A resting-state functional magnetic resonance imaging study with sibling design. Psychological Medicine 52, 1736–1745. doi:10.1017/S0033291720003529

Droutman, V., Bechara, A., Read, S. J., 2015. Roles of the Different Sub-Regions of the Insular Cortex in Various Phases of the Decision-Making Process. Front Behav Neurosci 9. doi: 10.3389/fnbeh.2015.00309

Dupont, G., van Rooij, D., Buitelaar, J.K., Reif, A., Grimm, O., 2022. Sex-related differences in adult attention-deficit hyperactivity disorder patients – An analysis of external globus pallidus functional connectivity in resting-state functional MRI. Front Psychiatry 13. 10.3389/fpsyt.2022.962911

Franke, B., Michelini, G., Asherson, P., Banaschewski, T., Bilbow, A., Buitelaar, J.K., Cormand, B., Faraone, S. V, Ginsberg, Y., Haavik, J., Kuntsi, J., Larsson, H., Lesch, K.-P., Ramos-Quiroga, J.A., Réthelyi, J.M., Ribases, M., Reif, A., 2018. Live fast, die young? A review on the developmental trajectories of ADHD across the lifespan. Eur Neuropsychopharmacol 28, 1059– 1088. 10.1016/j.euroneuro.2018.08.001

Gogolla, N., 2017. The insular cortex. Current Biology 27, R580–R586. 10.1016/j.cub.2017.05.010

Grimm, O., van Rooij, D., Hoogman, M., Klein, M., Buitelaar, J., Franke, B., Reif, A., Plichta, M.M., 2021. Transdiagnostic neuroimaging of reward system phenotypes in ADHD and comorbid disorders. Neurosci Biobehav Rev 128, 165–181. 10.1016/J.NEUBIOREV.2021.06.025

Hamzehpour, L., Bohn, T., Jaspers, L., Grimm, O., 2023. Exploring the link between functional connectivity of ventral tegmental area and physical fitness in schizophrenia and healthy controls. Eur Neuropsychopharmacol 76, 77–86. 10.1016/j.euroneuro.2023.07.009.

Hoogman, M., Bralten, J., Hibar, D.P., Mennes, M., Zwiers, M.P., Schweren, L.S.J., van Hulzen, K.J.E., Medland, S.E., Shumskaya, E., Jahanshad, N., Zeeuw, P. de, Szekely, E., Sudre, G., Wolfers, T., Onnink, A.M.H., Dammers, J.T., Mostert, J.C., Vives-Gilabert, Y., Kohls, G., Oberwelland, E., Seitz, J., Schulte-Rüther, M., Ambrosino, S., Doyle, A.E., Høvik, M.F., Dramsdahl, M., Tamm, L., van Erp, T.G.M., Dale, A., Schork, A., Conzelmann, A., Zierhut, K., Baur, R., McCarthy, H., Yoncheva, Y.N., Cubillo, A., Chantiluke, K., Mehta, M.A., Paloyelis, Y., Hohmann, S., Baumeister, S., Bramati, I., Mattos, P., Tovar-Moll, F., Douglas, P., Banaschewski, T., Brandeis, D., Kuntsi, J., Asherson, P., Rubia, K., Kelly, C., Martino, A. Di, Milham, M.P., Castellanos, F.X., Frodl, T., Zentis, M., Lesch, K.P., Reif, A., Pauli, P., Jernigan, T.L., Haavik, J., Plessen, K.J., Lundervold, A.J., Hugdahl, K., Seidman, L.J., Biederman, J., Rommelse, N., Heslenfeld, D.J., Hartman, C.A., Hoekstra, P.J., Oosterlaan, J., Polier, G. von, Konrad, K., Vilarroya, O., Ramos-Quiroga, J.A., Soliva, J.C., Durston, S., Buitelaar, J.K., Faraone, S. V., Shaw, P., Thompson, P.M., Franke, B., 2017. Subcortical brain volume differences in participants with attention deficit hyperactivity disorder in children and adults: a cross-sectional mega-analysis. Lancet Psychiatry 4, 310–319. 10.1016/S2215-0366(17)30049-4

Koob, G.F., Le Moal, M., 2008. Addiction and the Brain Antireward System. Annu Rev Psychol 59, 29–53. 10.1146/annurev.psych.59.103006.093548

Maier, S., Philipsen, A., Perlov, E., Runge, K., Matthies, S., Ebert, D., Endres, D., Domschke, K., Tebartz van Elst, L., Nickel, K., 2023. Left insular cortical thinning differentiates the inattentive and combined subtype of adult attention-deficit/hyperactivity disorder. J Psychiatr Res 159, 196–204. 10.1016/J.JPSYCHIRES.2023.01.030

Marek, S., Tervo-Clemmens, B., Calabro, F.J., Montez, D.F., Kay, B.P., Hatoum, A.S., Donohue, M.R., Foran, W., Miller, R.L., Hendrickson, T.J., Malone, S.M., Kandala, S., Feczko, E., Miranda-Dominguez, O., Graham, A.M., Earl, E.A., Perrone, A.J., Cordova, M., Doyle, O., Moore, L.A., Conan, G.M., Uriarte, J., Snider, K., Lynch, B.J., Wilgenbusch, J.C., Pengo, T., Tam, A., Chen, J., Newbold, D.J., Zheng, A., Seider, N.A., Van, A.N., Metoki, A., Chauvin, R.J., Laumann, T.O., Greene, D.J., Petersen, S.E., Garavan, H., Thompson, W.K., Nichols, T.E., Yeo, B.T.T., Barch, D.M., Luna, B., Fair, D.A., Dosenbach, N.U.F., 2022. Reproducible brain-wide association studies require thousands of individuals. Nature 603, 654–660. 10.1038/s41586-022-04492-9

Matthies, S., Sadohara-Bannwarth, C., Lehnhart, S., Schulte-Maeter, J., & Philipsen, A., 2018. The Impact of Depressive Symptoms and Traumatic Experiences on Quality of Life in Adults With ADHD. Journal of Attention Disorders, 22(5), 486–496. 10.1177/1087054716654568

Menon, V., 2011. Large-scale brain networks and psychopathology: a unifying triple network model. Trends Cogn Sci 15, 483–506. 10.1016/j.tics.2011.08.003

Menon, V., Uddin, L.Q., 2010. Saliency, switching, attention and control: a network model of insula function. Brain Struct Funct 214, 655–667. 10.1007/s00429-010-0262-0

Pauli, W.M., Nili, A.N., Tyszka, J.M., 2018. A high-resolution probabilistic in vivo atlas of human subcortical brain nuclei. Sci Data 5, 180063. 10.1038/sdata.2018.63

Picon, F.A., Sato, J.R., Anés, M., Vedolin, L.M., Mazzola, A.A., Valentini, B.B., Cupertino, R.B., Karam, R.G., Victor, M.M., Breda, V., Silva, K., da Silva, N., Bau, C.H.D., Grevet, E.H., Rohde, L.A.P., 2020. Methylphenidate Alters Functional Connectivity of Default Mode Network in Drug-Naive Male Adults With ADHD. J Atten Disord 24, 447–455. 10.1177/1087054718816822

Plichta, M.M., Scheres, A., 2014. Ventral–striatal responsiveness during reward anticipation in ADHD and its relation to trait impulsivity in the healthy population: A meta-analytic review of the fMRI literature. Neurosci Biobehav Rev 38, 125–134. 10.1016/j.neubiorev.2013.07.012

Rösler, M., Petra Retz-Junginger, P., Retz, W., Stieglitz, R.-D., 2021. HASE – Homburger ADHS-Skalen für Erwachsene. Hogrefe, Göttingen.

Seeley, W.W., Menon, V., Schatzberg, A.F., Keller, J., Glover, G.H., Kenna, H., Reiss, A.L., Greicius, M.D., 2007. Dissociable Intrinsic Connectivity Networks for Salience Processing and Executive Control. The Journal of Neuroscience 27, 2349–2356. 10.1523/JNEUROSCI.5587-06.2007

Shaw, P., De Rossi, P., Watson, B., Wharton, A., Greenstein, D., Raznahan, A., Sharp, W., Lerch, J.P., Chakravarty, M.M., 2014. Mapping the Development of the Basal Ganglia in Children With Attention-Deficit/Hyperactivity Disorder. J Am Acad Child Adolesc Psychiatry 53, 780–789.e11. 10.1016/J.JAAC.2014.05.003

Sheline, Y.I., Barch, D.M., Price, J.L., Rundle, M.M., Vaishnavi, S.N., Snyder, A.Z., Mintun, M.A., Wang, S., Coalson, R.S., Raichle, M.E., 2009. The default mode network and self-referential processes in depression. Proceedings of the National Academy of Sciences 106, 1942–1947. 10.1073/pnas.0812686106

Smallwood, J., Bernhardt, B.C., Leech, R., Bzdok, D., Jefferies, E., Margulies, D.S., 2021. The default mode network in cognition: a topographical perspective. Nat Rev Neurosci 22, 503–513. 10.1038/s41583-021-00474-4

Soman, S.M., Vijayakumar, N., Ball, G., Hyde, C., Silk, T.J., 2023. Longitudinal Changes of Resting-State Networks in Children With Attention-Deficit/Hyperactivity Disorder and Typically Developing Children. Biol Psychiatry Cogn Neurosci Neuroimaging 8, 514–521. 10.1016/j.bpsc.2022.01.001

Sonuga-Barke, E.J.S., 2005. Causal Models of Attention-Deficit/Hyperactivity Disorder: From Common Simple Deficits to Multiple Developmental Pathways. Biol Psychiatry 57, 1231–1238. 10.1016/J.BIOPSYCH.2004.09.008

Spencer, T.J., Biederman, J., Mick, E., 2007. Attention-Deficit/Hyperactivity Disorder: Diagnosis, Lifespan, Comorbidities, and Neurobiology. Ambulatory Pediatrics 7, 73–81. 10.1016/J.AMBP.2006.07.006

Stark, R., Bauer, E., Merz, C.J., Zimmermann, M., Reuter, M., Plichta, M.M., Kirsch, P., Lesch, K.P., Fallgatter, A.J., Vaitl, D., Herrmann, M.J., 2011. ADHD related behaviors are associated with brain activation in the reward system. Neuropsychologia 49, 426–434. 10.1016/j.neuropsychologia.2010.12.012

Sutcubasi, B., Metin, B., Kurban, M.K., Metin, Z.E., Beser, B., Sonuga-Barke, E., 2020. Resting-state network dysconnectivity in ADHD: A system-neuroscience-based meta-analysis. The World Journal of Biological Psychiatry 21, 662–672. 10.1080/15622975.2020.1775889

Tomasi, D., Volkow, N.D., 2012. Abnormal Functional Connectivity in Children with Attention-Deficit/Hyperactivity Disorder. Biol Psychiatry 71, 443–450. 10.1016/j.biopsych.2011.11.003

Uddin, L.Q., Kelly, A.M.C., Biswal, B.B., Margulies, D.S., Shehzad, Z., Shaw, D., Ghaffari, M., Rotrosen, J., Adler, L.A., Castellanos, F.X., Milham, M.P., 2008. Network homogeneity reveals decreased integrity of default-mode network in ADHD. J Neurosci Methods 169, 249–254. 10.1016/j.jneumeth.2007.11.031

Volkow, N.D., Wang, G.-J., Newcorn, J., Telang, F., Solanto, M. V., Fowler, J.S., Logan, J., Ma, Y., Schulz, K., Pradhan, K., Wong, C., Swanson, J.M., 2007. Depressed Dopamine Activity in Caudate and Preliminary Evidence of Limbic Involvement in Adults With Attention-Deficit/Hyperactivity Disorder. Arch Gen Psychiatry 64, 932. 10.1001/archpsyc.64.8.932

von Rhein, D., Cools, R., Zwiers, M.P., van der Schaaf, M., Franke, B., Luman, M., Oosterlaan, J., Heslenfeld, D.J., Hoekstra, P.J., Hartman, C.A., Faraone, S. V., van Rooij, D., van Dongen, E. V., Lojowska, M., Mennes, M., Buitelaar, J., 2015. Increased Neural Responses to Reward in Adolescents and Young Adults With Attention-Deficit/Hyperactivity Disorder and Their Unaffected Siblings. J Am Acad Child Adolesc Psychiatry 54, 394–402. 10.1016/j.jaac.2015.02.012

Wang, B., Wang, G., Wang, X., Cao, R., Xiang, J., Yan, T., Li, H., Yoshimura, S., Toichi, M., Zhao, S., 2021. Rich-Club Analysis in Adults With ADHD Connectomes Reveals an Abnormal Structural Core Network. J Atten Disord 25, 1068–1079. 10.1177/1087054719883031

Weibel, S., Menard, O., Ionita, A., Boumendjel, M., Cabelguen, C., Kraemer, C., Micoulaud-Franchi, J.A., Bioulac, S., Perroud, N., Sauvaget, A., Carton, L., Gachet, M., Lopez, R., 2020. Practical considerations for the evaluation and management of attention deficit hyperactivity disorder (ADHD) in adults. Encephale 46, 30–40. 10.1016/j.encep.2019.06.005

Wong, C.G., Stevens, M.C., 2012. The Effects of Stimulant Medication on Working Memory Functional Connectivity in Attention-Deficit/Hyperactivity Disorder. Biol Psychiatry 71, 458–466. 10.1016/j.biopsych.2011.11.011

Yankouskaya, A., Denholm-Smith, T., Yi, D., Greenshaw, A.J., Cao, B., Sui, J., 2022. Neural Connectivity Underlying Reward and Emotion-Related Processing: Evidence from a Large-Scale Network Analysis. Front Syst Neurosci 16, 833625. 10.3389/fnsys.2022.833625

